# Influenza vaccine effectiveness against influenza A-associated hospitalization and severe in-hospital outcomes among adults in the United States, 2024–2025

**DOI:** 10.64898/2026.03.31.26349873

**Authors:** Nathaniel M. Lewis, Seana Cleary, Elizabeth J. Harker, Basmah Safdar, Adit A. Ginde, Ithan D. Peltan, Manjusha Gaglani, Cristie Columbus, Emily T. Martin, Adam S. Lauring, Jay S. Steingrub, David N. Hager, Amira Mohamed, Nicholas J Johnson, Akram Khan, Abhijit Duggal, Jennifer G. Wilson, Nida Qadir, Laurence W. Busse, Jennie H. Kwon, Matthew C. Exline, Ivana A. Vaughn, Jarrod M Mosier, Estelle S. Harris, Yuwei Zhu, Carlos G. Grijalva, Natasha B. Halasa, James Chappell, Diya Surie, Fatimah S Dawood, Sascha Ellington, Wesley H Self, the Investigating Viruses in the Acutely Ill (IVY) Network

**Affiliations:** Influenza Division, National Center for Immunization and Respiratory Diseases, Centers for Disease Control and Prevention, Atlanta, Georgia, USA; STI Federal, Sault Tribe Incorporated, Sault Ste Marie, Michigan, USA; Department of Emergency Medicine, Yale University, New Haven, Connecticut, USA; Department of Emergency Medicine, University of Colorado, Aurora, Colorado, USA; Department of Medicine, Intermountain Medical Center, Murray, Utah, USA; Department of Pediatrics, Baylor Scott & White Health, Temple, Texas, USA; College of Medicine, Baylor University, Temple, Texas, USA; Department of Medicine, Baylor Scott & White Health, Dallas, Texas, USA; Department of Medicine, Texas A&M University College of Medicine, Dallas, Texas, USA; School of Public Health, University of Michigan, Ann Arbor, Michigan, USA; Department of Medicine, University of Michigan, Ann Arbor, Michigan, USA; Department of Medicine, Baystate Medical Center, Springfield, Massachusetts, USA; Department of Medicine, Johns Hopkins University School of Medicine, Baltimore, Maryland, USA; Department of Medicine, Montefiore Medical Center, Bronx, New York, USA; Department of Emergency Medicine, University of Washington, Seattle, Washington, USA; Department of Medicine, Oregon Health and Science University, Portland, Oregon, USA; Department of Medicine, Cleveland Clinic, Cleveland, Ohio, USA; Department of Emergency Medicine, Stanford University, Stanford, California, USA; Department of Medicine, University of California, Los Angeles, California, USA; Department of Medicine, Emory University, Atlanta, Georgia, USA; Department of Medicine, Northwestern University, Chicago, Illinois, USA; Department of Medicine, The Ohio State University, Columbus, Ohio, USA; Department of Public Health Sciences, Henry Ford Health, Detroit, Michigan, USA; Department of Emergency Medicine, University of Arizona, Tucson, Arizona, USA; Department of Medicine, University of Utah, Salt Lake City, Utah, USA; Department of Biostatistics, Vanderbilt University Medical Center, Nashville, Tennessee, USA; Department of Health Policy, Vanderbilt University Medical Center, Nashville, Tennessee, USA; Department of Pediatrics, Vanderbilt University Medical Center, Nashville, Tennessee, USA; Coronavirus and Other Respiratory Viruses Division, CDC, Atlanta, Georgia, USA; Vanderbilt Institute for Clinical & Translational Research, Vanderbilt University Medical Center, Nashville, Tennessee, USA

**Author notes:** Corresponding author: Sascha Ellington, Influenza Division, National Center for Immunization and Respiratory Diseases, Centers for Disease Control and Prevention, 1600 Clifton Road, Atlanta, Georgia, USA, 30329,.

## Abstract

**Background:** The U.S. 2024–2025 influenza season was characterized by sustained elevated activity from November 2024–April 2025, with circulation of both influenza A(H1N1)pdm09 and A(H3N2), the latter of which included some antigenically drifted viruses.

**Methods:** From October 1, 2024, to April 30, 2025, a multistate respiratory virus surveillance network enrolled adults hospitalized with acute respiratory illness in 26 U.S. medical centers. Influenza vaccine effectiveness (VE) against influenza-associated hospitalization and severe in-hospital outcomes was estimated using a test-negative study. The odds of influenza vaccination among influenza-positive case patients and influenza-negative control patients were compared using multivariable logistic regression; VE was calculated as (1 − adjusted odds ratio for vaccination) × 100, expressed as a percent.

**Results:** The 2024–2025 seasonal influenza vaccine was effective against influenza-associated hospitalization (VE: 40% [95% confidence interval (CI): 32%–47%]), consistent across age group and influenza A subtypes. Influenza vaccination also reduced the overall risk of all severe in-hospital outcomes evaluated, including standard oxygen therapy (VE: 41% [95% CI: 31%–50%]), non-invasive advanced respiratory support (VE: 38% [95% CI: 19%–52%]), invasive organ support (VE: 58% [95% CI: 44%–69%]), ICU admission (VE: 58% [95% CI: 47%–67%]), and death (VE: 52% [95% CI: 18%–71%]) with effectiveness varying by influenza A subtype and age.

**Conclusions:** Influenza vaccination reduced the risk of influenza-related hospitalization and severe in-hospital outcomes in adults during the severe 2024–2025 influenza season compared to those not vaccinated.

## Introduction

The U.S. 2024–2025 influenza season was characterized by sustained elevated activity from November 2024 through April 2025, peaking during the last week of January 2025 and resulting in the most severe season recorded since before the COVID-19 pandemic. There was roughly equal circulation of influenza A(H1N1)pdm09 and A(H3N2) throughout the season, with minimal circulation of influenza B.^1^

The 2024–2025 Northern Hemisphere influenza vaccines, several components remained unchanged from the 2023–2024 vaccines, including an A/Victoria/4897/2022 A(H1N1)pdm09-like virus and a B/Austria/135917/2021 (B/Victoria lineage)-like virus in the egg-based vaccine and an A/Wisconsin/67/2022 A(H1N1)pdm09-like virus in both the cell- and recombinant-based vaccines. The B/Yamagata components were removed from all vaccines due to the absence of detection of this strain after March 2020. Finally, the A(H3N2) component was changed from an A/Darwin/9/2021 to an A/Thailand/8/2022 (H3N2)-like virus in the egg-based vaccine and from an A/Darwin/6/2021 (H3N2)-like virus to an A/Massachusetts/18/2022 (H3N2)-like virus in the cell- and recombinant-based vaccines. Genetic and antigenic characterization of circulating viruses in the U.S. showed that while most circulating A(H1N1)pdm09 and B viruses were antigenically similar to vaccine reference viruses, some A(H3N2) viruses were antigenically drifted from the vaccine virus.^2^

Reduced influenza vaccine effectiveness (VE) against influenza A(H3N2) is now well documented and can arise from egg-adapted changes to vaccine viruses during vaccine production, antigenic drift of circulating viruses either pre- or intra-seasonally from the vaccine strain, as well as limited early-in-life exposure to the virus among adults born before 1968 when A(H3N2) viruses first began circulating.^3–6^ Less, however, is known about whether VE differs against severe outcomes associated with influenza A(H3N2) compared with A(H1N1)pdm09. Recent systematic reviews comparing VE by virus type and subtype have shown reduced VE against both influenza A(H3N2) and a range of A(H3N2)-associated severe outcomes (including hospitalization, pneumonia, and intensive care unit [ICU] admission, especially among older adults and during influenza seasons with antigenic drift of A(H3N2) viruses.^6–8^ However, some reviews have shown protection against A(H3N2)-associated mortality.^7,8^ The 2024-2025 influenza season was severe with roughly equal circulation of influenza A(H1N1)pdm09 and A(H3N2) viruses, providing an opportunity to study differences in VE against severe in-hospital outcomes associated with the two influenza A subtypes.

## Methods

### Surveillance Network

The Investigating Respiratory Viruses in the Acutely Ill (IVY) Network, comprised of 26 hospitals across 20 U.S. states, was a surveillance network primarily used to estimate VE against influenza-, coronavirus disease 2019 (COVID-19)-, and respiratory syncytial virus (RSV)-associated hospitalization in adults from 2019–2025.^9^ This program was reviewed by CDC and each participating site, deemed not research by CDC, and was conducted in accordance with applicable federal law and CDC policy^§^. This analysis includes enrolled patients hospitalized within the IVY Network from October 1, 2024, to April 30, 2025. Written informed consent was obtained from each patient or a legally authorized representative.

### Study Population

Each site assessed eligibility and collected data and specimens according to standard protocols (see Supplementary Materials). During the 2024–2025 influenza season, sites recruited hospitalized adults ≥18 years who met an acute respiratory illness (ARI) case definition (≥1 of the following: fever, cough, shortness of breath, new hypoxemia, new pulmonary findings on chest imaging consistent with pneumonia) and received a molecular or antigen influenza test. Case patients were those who tested positive for influenza virus by local clinical testing and/or centralized reverse transcription polymerase chain reaction (RT-PCR) testing. Cases with SARS-CoV-2 or RSV coinfections were excluded from analyses. Control patients were those who tested negative for influenza by both local clinical testing and RT-PCR. Controls that tested positive for SARS-CoV-2 (patients aged ≥18 years) and/or RSV (adults aged ≥60 years) were excluded to minimize confounding bias from potentially correlated vaccination status across pathogens.^10,11^ Controls were enrolled at the same hospital within two weeks of enrollment in a roughly 1:1 case-control ratio without matching.

### Centralized Viral Testing

Sites completed clinical viral testing in the hospitals and shipped nasal swab specimens to Vanderbilt University Medical Center (VUMC; Nashville, TN) to undergo RT-PCR testing for influenza viruses, SARS-CoV-2, and RSV (see Supplementary Materials). Specimens that tested positive for influenza virus by RT-PCR then underwent viral whole-genome sequencing (WGS) at the University of Michigan (Ann Arbor, MI).

### Influenza Vaccination History

Influenza vaccination status was determined using electronic medical records, immunization information system (IIS) searches, and self- or surrogate-report that included a plausible date and location of vaccine administration. Patients were considered vaccinated if they received at least one dose of the 2024–2025 seasonal influenza vaccine ≥14 days before illness onset. Patients were considered unvaccinated if they received no doses of the 2024–2025 seasonal influenza vaccine prior to illness onset.

### Exclusion Criteria

Patients were additionally excluded if they: (1) received no viral testing <10 days after illness onset date, (2) received their first viral testing after illness onset and >3 days after hospital admission, (3) had an illness onset date after hospital admission or ≥14 days before hospital admission, (4) were enrolled as a control before the first influenza case or after the last influenza case at a local site, (5) had an unknown influenza vaccination status, or (6) received an influenza vaccination <14 days before illness-onset date.

### Statistical Analysis

We compared patient characteristics by case status using Chi-square tests for categorical variables and Mann-Whitney U tests for continuous variables. We used a test-negative design to estimate influenza VE. Our primary outcome was influenza-associated hospitalization, defined as hospital admission for symptomatic, laboratory-confirmed influenza virus infection. We first fit a multivariable logistic regression model to compare the odds of influenza vaccination among cases and controls, adjusting for age group (18–49, 50–64, ≥65 years), sex (male/female), race/ethnicity (non-Hispanic White, non-Hispanic Black, Hispanic, non-Hispanic other, other), Census region (Northeast, Midwest, South, West), and month and year of enrollment. Additional covariates, including chronic respiratory condition, immunocompetence status, and smoking status, were considered for inclusion in the model but ultimately not retained as their inclusion did not change VE by ≥5%. If the model did not converge well due to separation, we fit a Firth penalized regression model using the same covariables. We then calculated VE for protection against influenza-associated hospitalization as (1 – adjusted odds ratio for vaccination) × 100, expressed as a percent. We calculated overall VE and stratified by influenza A virus subtype, age group, immunocompetence status, and underlying conditions. When stratifying by age group, age group was not included as a covariable. We calculated VE by time from vaccination to illness onset using four exposure levels (unvaccinated and vaccinated 14–59, 60–119, and >120 days prior to illness onset).

In secondary analyses, we further assessed VE against hospitalization with severe in-hospital influenza-associated outcomes using the same covariables as used in the primary analysis. Cases were restricted to influenza-positive patients who met each of the outcome definitions, and controls remained hospitalized patients who tested negative for influenza virus. In-hospital outcomes included: (1) use of standard supplemental oxygen therapy (≤30 L/min, excluding patients using supplemental oxygen chronically prior to admission), (2) use of non-invasive advanced respiratory support (including high-flow nasal cannula or non-invasive ventilation, and excluding patients using advanced respiratory support chronically prior to admission), (3) invasive organ support (including new invasive mechanical ventilation, renal replacement therapy, or vasopressor use), (4) ICU admission, and (5) death (see Supplementary Materials). We stratified these models by influenza A subtype and age group. A *p*-value of <0.05 or a 95% confidence interval (CI) excluding the null value were considered significant for all analyses. Analyses were performed using R version 4.4.1 (R Core Team, Vienna, Austria)^12^ and SAS version 9.4 (Cary, North Carolina, USA).^13^

## Results

### Enrollment and Patient Characteristics

Sites enrolled 10,252 patients between October 1, 2024, and April 30, 2025. A total of 4,143 patients were excluded from the analysis, most commonly for testing positive for SARS-CoV-2 or RSV (n=2,375; 57%) or being enrolled as a control before the first influenza case or after the last influenza case at a local site (n=1,233; 30%; Figure 1). The final analytic dataset was comprised of 6,109 patients, including 1,735 influenza cases and 4,374 controls.

**Figure 1.**
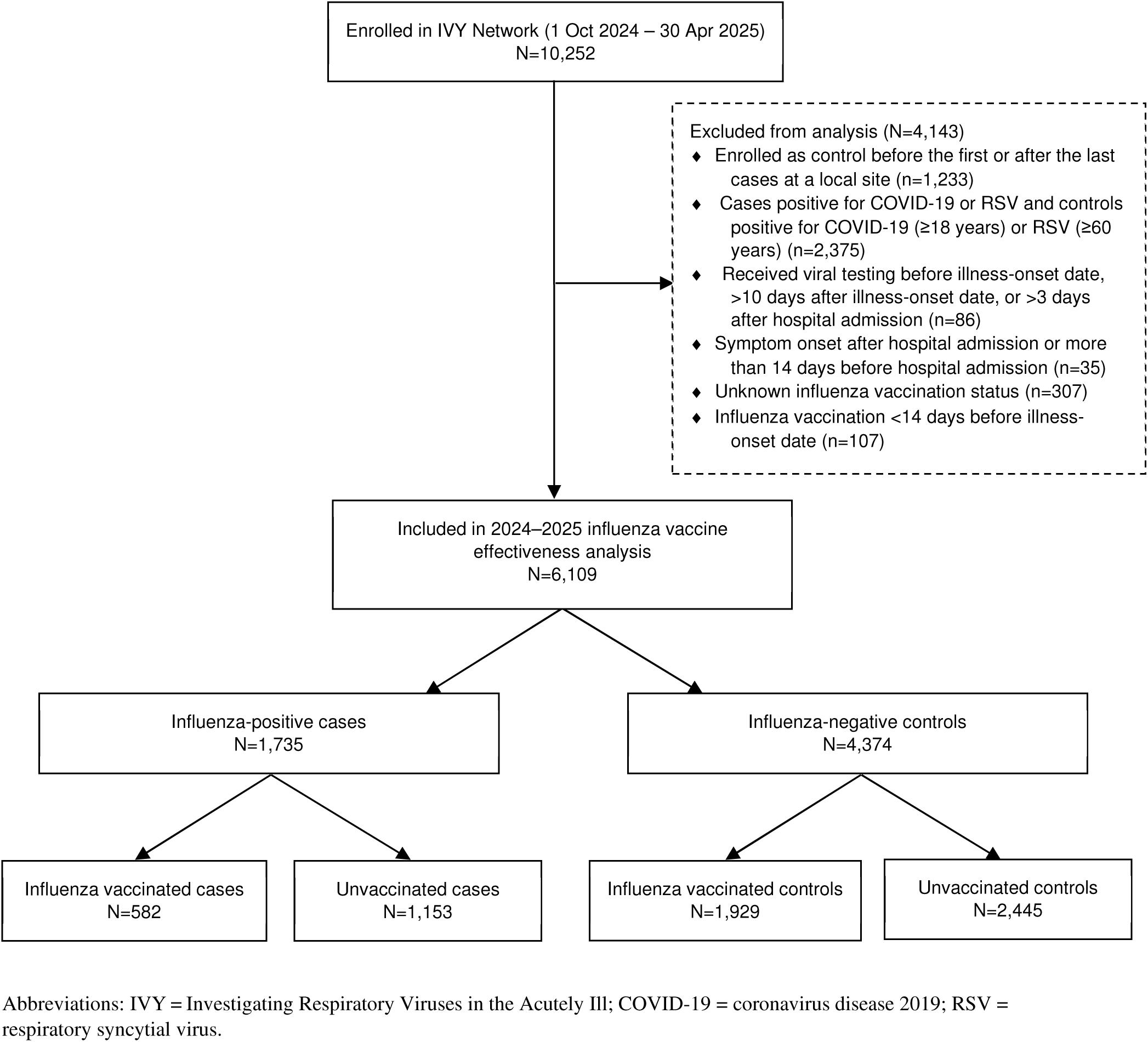
Exclusion flowchart, IVY Network, 2024–2025.

Influenza virus type/subtype was determined by RT-PCR for 1,113 cases. Of these, 647 (58%) had influenza A(H1N1)pdm09 and 466 (42%) had influenza A(H3N2); none had influenza B. Among those with influenza A(H1N1)pdm09, a genetic subclade was identified by viral WGS in 309 patients, including 186 (60%) belonging to 6B.1A.5a.2a.1 and 123 (40%) belonging to 6B.1A.5a.2a. Among those with influenza A(H3N2), a subclade was identified by WGS in 249 patients, all of which were 3C.2a1b.2a.2a.3a.1.

The percentage of the population who received influenza vaccination was lower among influenza case patients than control patients (34% vs 44%; p<0.01; Table 1). Influenza case patients, compared with control patients, were more likely to smoke (25% vs 21%; p<0.01) and less likely to be immunocompromised (21% vs 29%; p<0.01), to have a chronic respiratory condition (40% vs 44%, p=0.01), and be hospitalized in the previous year (50% vs 66%; p<0.01). Patients who received an influenza vaccine, compared to unvaccinated patients, were older (median: 69 vs 62 years; p<0.01), more often non-Hispanic White (61% vs 51%; p<0.01), more likely to have health insurance (99% vs 95%; p<0.01), and less likely to smoke (15% vs 27%; p<0.01). They also were more likely to be immunocompromised (30% vs 24%; p<0.01), have a chronic respiratory condition (47% vs 40%; p<0.01), and to have been hospitalized in the previous year (66% vs 58%; p<0.01; see Supplementary Table 1). Among patients aged 18–64 years with documentation on influenza vaccine product received, most received the standard-dose egg-based inactivated (n=543; 73%), standard-dose cell culture-based inactivated (n=94; 13%), or recombinant influenza vaccine (n=51; 7%). Among patients aged ≥65 years with documentation on influenza vaccine product received, most received the high-dose egg-based inactivated influenza vaccine (n=825; 62%) or standard-dose egg-based inactivated influenza vaccine with an MF59 adjuvant (n=419; 32%).

**Table 1.**
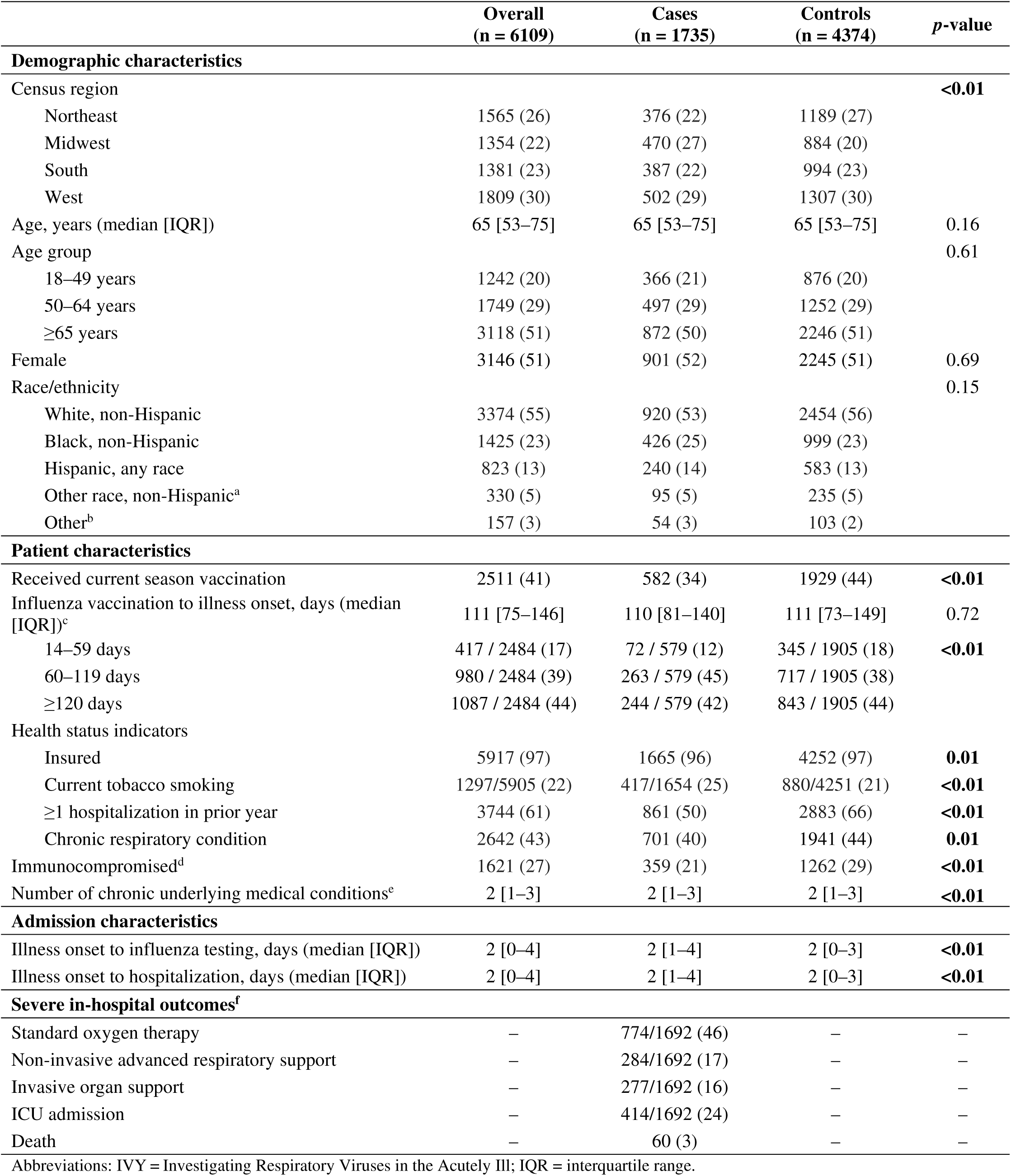
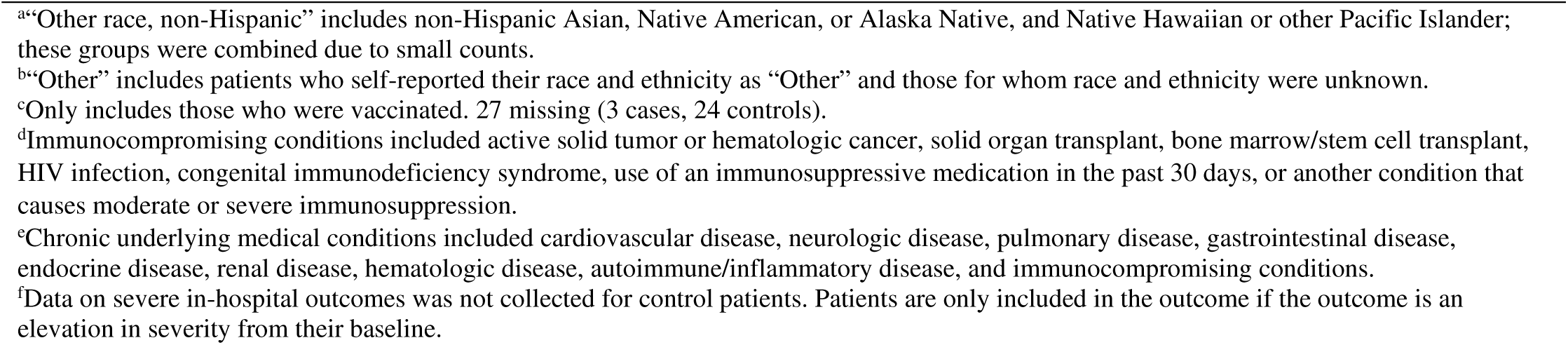
Demographic and clinical characteristics of hospitalized influenza cases and controls, IVY Network, 2024–2025.

|  | **Overall**<br>**(n = 6109)** | **Cases**<br>**(n = 1735)** | **Controls**<br>**(n = 4374)** | **p-value** |
| --- | --- | --- | --- | --- |
| **Demographic characteristics** |  |  |  |  |
| Census region |  |  |  | **<0.01** |
| Northeast | 1565 (26) | 376 (22) | 1189 (27) |  |
| Midwest | 1354 (22) | 470 (27) | 884 (20) |  |
| South | 1381 (23) | 387 (22) | 994 (23) |  |
| West | 1809 (30) | 502 (29) | 1307 (30) |  |
| Age, years (median [IQR]) | 65 [53–75] | 65 [53–75] | 65 [53–75] | 0.16 |
| Age group |  |  |  | 0.61 |
| 18–49 years | 1242 (20) | 366 (21) | 876 (20) |  |
| 50–64 years | 1749 (29) | 497 (29) | 1252 (29) |  |
| ≥65 years | 3118 (51) | 872 (50) | 2246 (51) |  |
| Female | 3146 (51) | 901 (52) | 2245 (51) | 0.69 |
| Race/ethnicity |  |  |  | 0.15 |
| White, non-Hispanic | 3374 (55) | 920 (53) | 2454 (56) |  |
| Black, non-Hispanic | 1425 (23) | 426 (25) | 999 (23) |  |
| Hispanic, any race | 823 (13) | 240 (14) | 583 (13) |  |
| Other race, non-Hispanic<sup>a</sup> | 330 (5) | 95 (5) | 235 (5) |  |
| Other<sup>b</sup> | 157 (3) | 54 (3) | 103 (2) |  |
| **Patient characteristics** |  |  |  |  |
| Received current season vaccination | 2511 (41) | 582 (34) | 1929 (44) | **<0.01** |
| Influenza vaccination to illness onset, days (median [IQR])<sup>c</sup> | 111 [75–146] | 110 [81–140] | 111 [73–149] | 0.72 |
| 14–59 days | 417 / 2484 (17) | 72 / 579 (12) | 345 / 1905 (18) | **<0.01** |
| 60–119 days | 980 / 2484 (39) | 263 / 579 (45) | 717 / 1905 (38) |  |
| ≥120 days | 1087 / 2484 (44) | 244 / 579 (42) | 843 / 1905 (44) |  |
| Health status indicators |  |  |  |  |
| Insured | 5917 (97) | 1665 (96) | 4252 (97) | **0.01** |
| Current tobacco smoking | 1297/5905 (22) | 417/1654 (25) | 880/4251 (21) | **<0.01** |
| ≥1 hospitalization in prior year | 3744 (61) | 861 (50) | 2883 (66) | **<0.01** |
| Chronic respiratory condition | 2642 (43) | 701 (40) | 1941 (44) | **0.01** |
| Immunocompromised<sup>d</sup> | 1621 (27) | 359 (21) | 1262 (29) | **<0.01** |
| Number of chronic underlying medical conditions<sup>e</sup> | 2 [1–3] | 2 [1–3] | 2 [1–3] | **<0.01** |
| **Admission characteristics** |  |  |  |  |
| Illness onset to influenza testing, days (median [IQR]) | 2 [0–4] | 2 [1–4] | 2 [0–3] | **<0.01** |
| Illness onset to hospitalization, days (median [IQR]) | 2 [0–4] | 2 [1–4] | 2 [0–3] | **<0.01** |
| **Severe in-hospital outcomes<sup>f</sup>** |  |  |  |  |
| Standard oxygen therapy | – | 774/1692 (46) | – | – |
| Non-invasive advanced respiratory support | – | 284/1692 (17) | – | – |
| Invasive organ support | – | 277/1692 (16) | – | – |
| ICU admission | – | 414/1692 (24) | – | – |
| Death | – | 60 (3) | – | – |

Abbreviations: IVY = Investigating Respiratory Viruses in the Acutely Ill; IQR = interquartile range.
<sup>a</sup>“Other race, non-Hispanic” includes non-Hispanic Asian, Native American, or Alaska Native, and Native Hawaiian or other Pacific Islander; these groups were combined due to small counts.
<sup>b</sup>“Other” includes patients who self-reported their race and ethnicity as “Other” and those for whom race and ethnicity were unknown.
<sup>c</sup>Only includes those who were vaccinated. 27 missing (3 cases, 24 controls).
<sup>d</sup>Immunocompromising conditions included active solid tumor or hematologic cancer, solid organ transplant, bone marrow/stem cell transplant, HIV infection, congenital immunodeficiency syndrome, use of an immunosuppressive medication in the past 30 days, or another condition that causes moderate or severe immunosuppression.
<sup>e</sup>Chronic underlying medical conditions included cardiovascular disease, neurologic disease, pulmonary disease, gastrointestinal disease, endocrine disease, renal disease, hematologic disease, autoimmune/inflammatory disease, and immunocompromising conditions.
<sup>f</sup>Data on severe in-hospital outcomes was not collected for control patients. Patients are only included in the outcome if the outcome is an elevation in severity from their baseline.

In the first 28 days of hospitalization, 774 (46%) influenza cases used standard oxygen therapy, 284 (17%) used non-invasive advanced respiratory support (of which 102 [36%] had high-flow nasal cannula and 182 [64%] had non-invasive ventilation), 277 (16%) received invasive organ support (of which 203 [73%] had invasive mechanical ventilation, 34 [12%] had renal replacement therapy and 226 [82%] had vasopressor use) , 414 (24%) were admitted to the ICU, and 60 (3%) died (Table 1).

### VE Against Influenza-Associated Hospitalization

The overall VE against influenza-associated hospitalization was 40% (95% CI: 32%–47%; Figure 2). Stratified by age group, VE was 50% (95% CI: 31%–63%) among adults aged 18–49 years, 41% (95% CI: 25%–53%) among adults aged 50–64 years, and 39% (28%–48%) among adults aged ≥65 years. By immunocompetence status, VE was 47% (95% CI: 39%–54%) for immunocompetent patients. All results were statistically significant with the exception of VE for immunocompromised patients (7% [95% CI: - 19%–28%]), where the confidence interval crossed zero. Immunocompetent patients with 0–3 underlying conditions had a VE of 48% (95% CI: 39%–56%), while those with ≥4 underlying conditions had a VE of 31% (95% CI: 3%–51%]). The VE against influenza-associated hospitalization was 50% (95% CI: 35%–62%) in those vaccinated 14–59 days prior to illness onset, 40% (95% CI: 29%–49%) in those vaccinated 60–119 days prior to illness onset, and 35% (95% CI: 23%–46%) in those vaccinated ≥120 days prior to illness onset.

**Figure 2.**
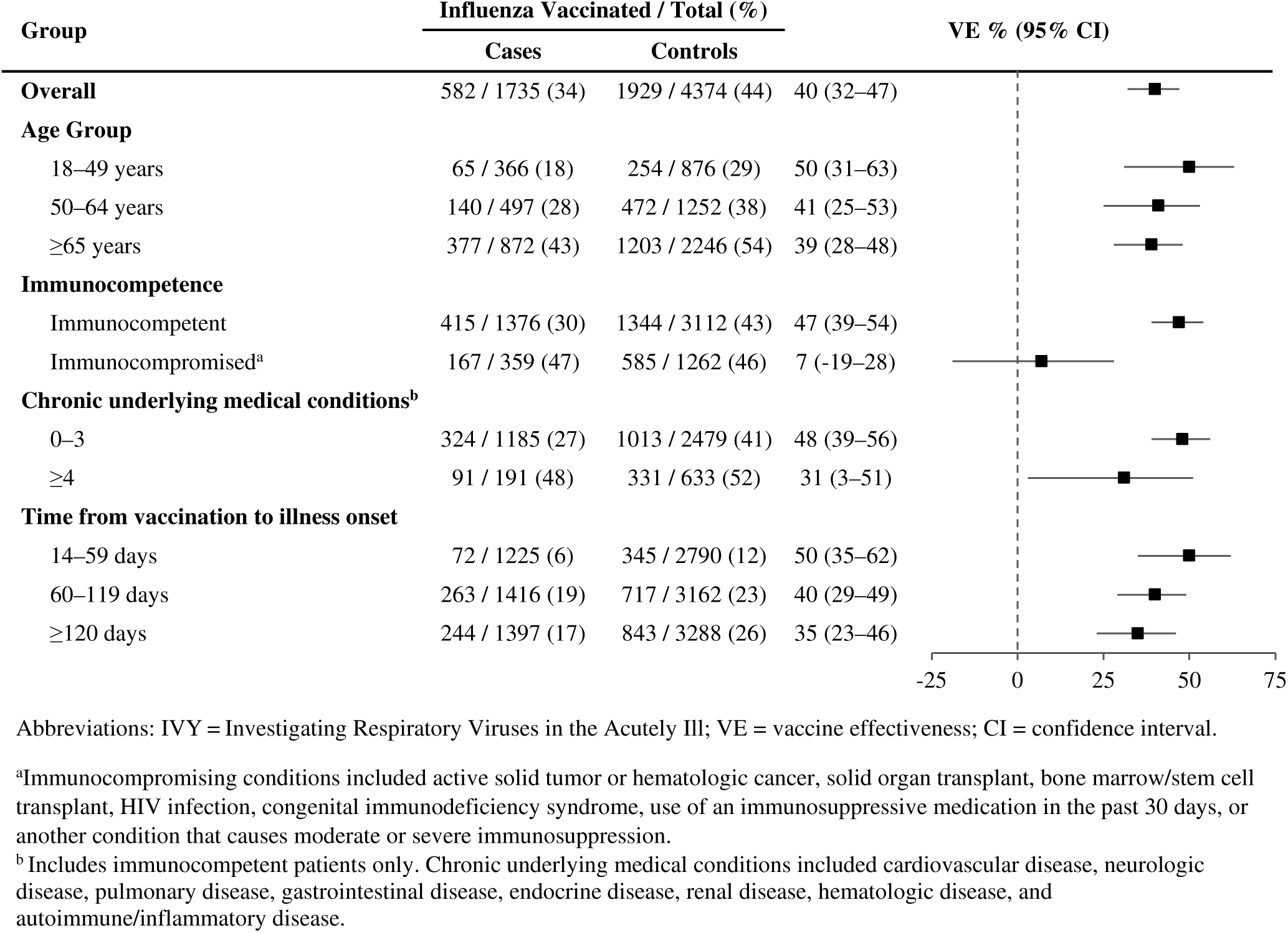
Influenza VE against hospitalization overall and by age group, immunocompetence status, and time since vaccination to illness onset; IVY Network, 2024–2025.

Influenza vaccination was protective against both influenza A(H1N1)pdm09-associated hospitalization (VE: 42% [95% CI: 30%–51%]) and influenza A(H3N2)-associated hospitalization (VE: 44% [95% CI: 30%–55%]; Figure 3). Among the A(H1N1)pdm09 subclades, VE against 6B.1A.5a.2a.1-associated hospitalization was 47% (95% CI: 27%–61%) and VE against 6B.1A.5a.2a-associated hospitalization was 45% (95% CI: 19%–63%). VE against influenza A(H1N1)pdm09-associated hospitalization was consistent across age groups 18–64 years (42% [95% CI: 23%–57%]) and ≥65 years (40% [95% CI: 24%–53%]). VE against influenza A(H3N2)-associated hospitalizations was 58% (95% CI: 39%–71%) for those 18–64 years old and 35% (95% CI: 15%–50%) for those ≥65 years old.

**Figure 3.**
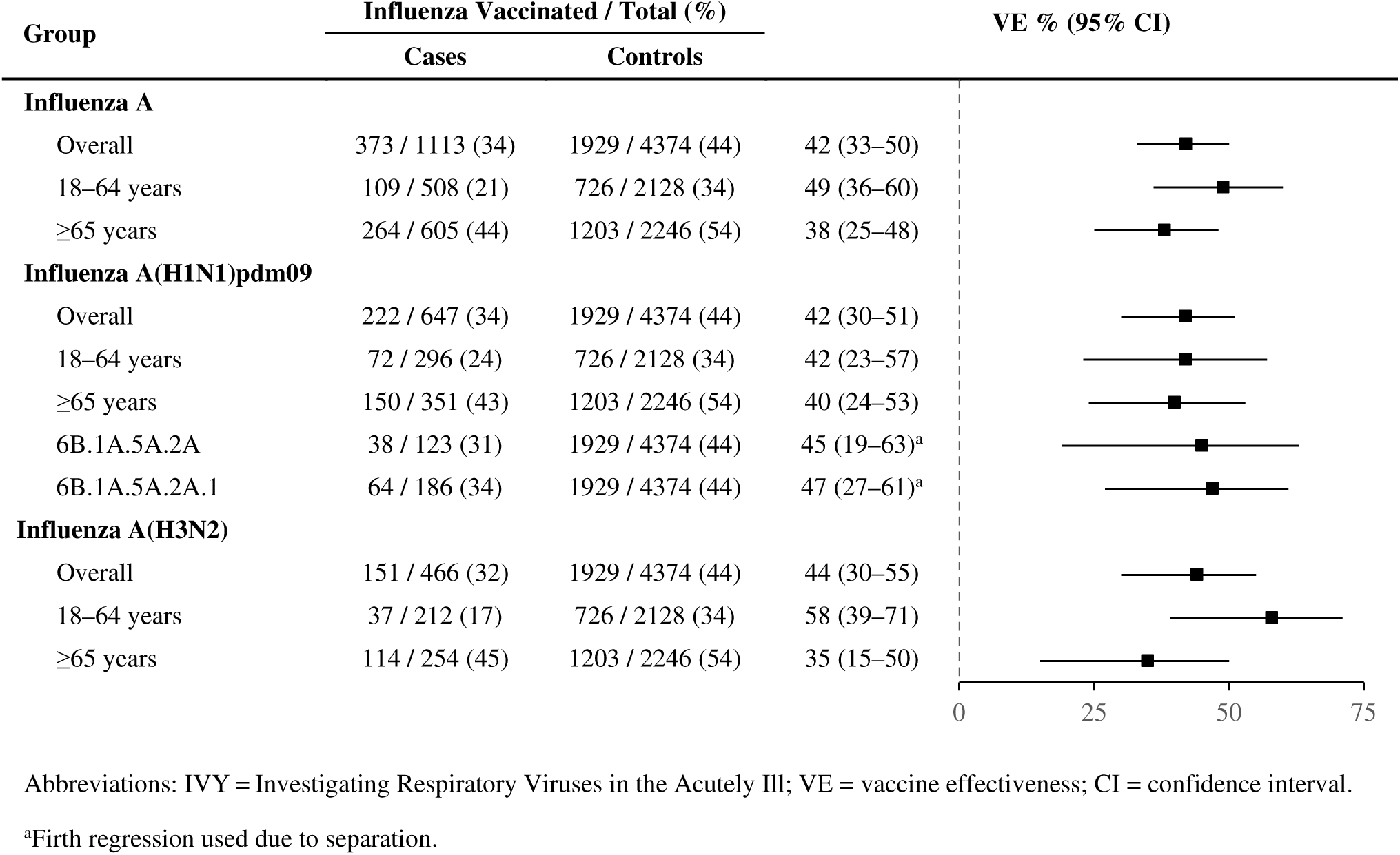
Influenza VE against hospitalization by influenza A type and subtype, stratified by age and subclade; IVY Network, 2024–2025.

### VE Against Influenza-Associated Severe In-Hospital Outcomes

Influenza vaccination was effective against all evaluated severe in-hospital outcomes, reducing the risk of the patient requiring use of standard oxygen therapy (41% [95% CI: 31%–50%]), non-invasive advanced respiratory support (38% [95% CI: 19%–52%]), invasive organ support (58% [95% CI: 44%–69%]), ICU admission (58% [95% CI: 47%–67%]), and death (52% [95% CI: 18%–71%]) (Figure 4).

**Figure 4.**
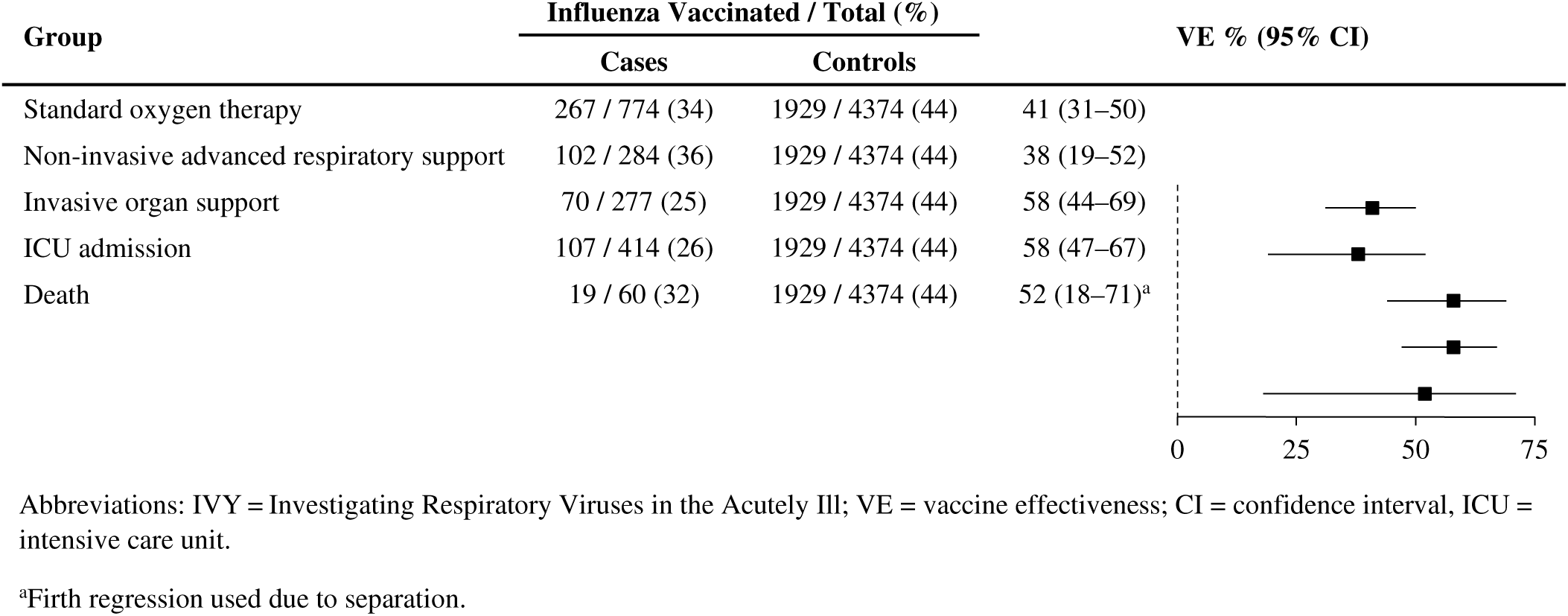
VE against influenza-associated severe in-hospital outcomes in hospitalized adults; IVY Network, 2024–2025.

When stratified by influenza A subtype, VE against influenza-associated hospitalization with use of non-invasive advanced respiratory support and hospitalization with death in those with influenza A(H3N2) crossed the null. VE against hospitalization with all other severe in-hospital outcomes remained significant when stratified by influenza A subtype (Figure 5), although VE estimates for all influenza A(H3N2)-associated outcomes (excepting standard oxygen therapy) were similar to but lower than those for the corresponding outcome associated with influenza A(H1N1)pdm09. When stratified by age group, VE against hospitalization with use of non-invasive advanced respiratory support in those aged 18–64 years and against hospitalization with progression to death in those ≥65 years crossed the null. VE against hospitalization with all other measured severe in-hospital outcomes remained significant when stratified by age group (see Supplementary Figure 1).

**Figure 5.**
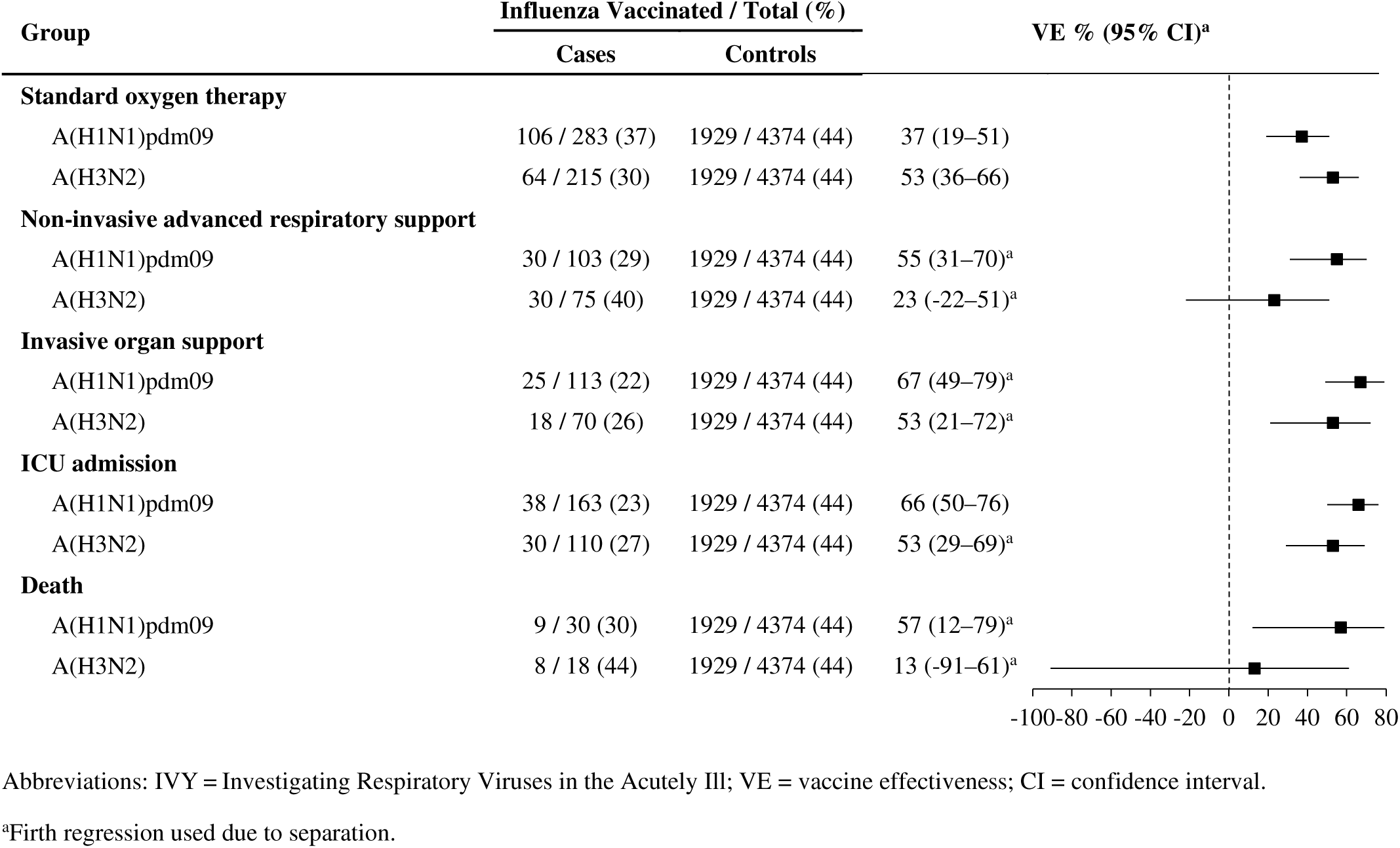
Influenza VE against influenza-associated severe in-hospital outcomes in hospitalized adults, stratified by influenza A subtype; IVY Network, 2024–2025.

## Discussion

In this analysis of influenza vaccine effectiveness during the 2024–2025 influenza season, we found consistent protection against influenza A(H1N1)pdm09- and A(H3N2)-associated hospitalization in adults. In addition, protection remained consistent at all intervals of time since vaccination, which may have contributed to an overall VE estimate similar to the 2023–2024 season in this network^3^ despite a longer period of sustained elevated activity during the 2024–2025 season. Our finding of >50% VE against influenza–associated mortality generally adds to the growing body of work demonstrating significant VE against death.^7,8,14,15^ In addition, our use of a test-negative VE study design addresses earlier concerns regarding potential overestimation of a mortality benefit due to unadjusted selection bias in previous cohort studies.^16^ Finally, our finding of 48% VE among patients with ≤3 categories of underlying conditions, mirrors previous studies showing substantial VE among patients with 0–2 conditions^17^, although our slightly lower estimate for patients with ≥4 conditions may suggest that a high burden of underlying conditions may affect VE. Although our finding of null protection among immunocompromised patients in this network diverges from the previous two seasons where significant protection was observed^3,18^, null protection was also observed during an A(H3N2)-dominant season with antigenic drift and potentially different makeup of a heterogeneous immunocompromised patient group that may change season to season.^19^ The absence of a measurable benefit in protection against hospitalization for immunocompromised patients supports the use of other strategies in this population in addition to vaccination, including early antiviral treatment to reduce severe outcomes upon admission.^20^

Influenza subtype and patient age both appeared to affect VE. Protection against A(H1N1)pdm09- and A(H3N2)-associated hospitalization was similar overall. However, patients aged ≥65 years experienced less protection from influenza vaccination compared with patients aged 18–64 years against hospitalization associated with influenza A(H3N2), similar to previous seasons in this network^3^ and others.^6^ In addition, estimates of protection against severe in-hospital outcomes diverged between subtypes, with similar but less consistent protection against influenza A(H3N2)-associated outcomes than A(H1N1)pdm09-associated outcomes. There was some reduction in VE point estimates against the three most severe outcomes (i.e., invasive organ support, ICU admission, and death) among patients aged ≥65 years, mirroring studies observing larger age-related gaps with more severe outcomes.^21,22^ However, other studies focused exclusively on VE against death among patients aged ≥65 years have found significant protection against influenza-associated death during prior seasons.^14,15^ Reduced protection against influenza A(H3N2) compared to A(H1N1)pdm09, especially among patients aged ≥65 years, has been observed consistently in recent studies.^5–7,21,23^ Although we generally found the same this season with regard to the most severe outcomes, this was not the case for hospitalization or non-invasive ventilation, mirroring other studies that found larger gaps between A(H1N1)pdm09 and A(H3N2) VE for more severe outcomes.^8,21,23^ It may therefore be that during a season when the influenza vaccine viruses and circulating viruses are well matched, A(H3N2) VE is reduced mostly among patients aged ≥65 years who lack early immune imprinting to A(H3N2) viruses^4,5^, and may be predisposed to worse influenza–associated outcomes, rather than due to antigenic drift that may affect all patients similarly and substantially affect protection against even less severe forms of illness.

Our results must be considered in light of several limitations. First, statistical power for some estimates of VE against in-hospital outcomes was limited. Reduced precision for rarer outcomes may explain why some studies have demonstrated that protection increases with the severity of outcome^14,21,24^ while others have not, particularly when more granular outcomes on the severity continuum are assessed.^8,18,22^ VE against progression, measured by comparing vaccination among cases with a specific influenza-associated outcome (e.g., ICU admission, death) to cases without the same outcome has been employed in studies in Catalonia (Spain)^25,26^ and may offer an alternative method to assessing effectiveness against severe outcomes. Relatedly, sample size limited the ability to conduct more sub-analyses for smaller populations (e.g., about differential VE against subtype-specific severe outcomes in different age groups). Future multi-season analyses are necessary to assess, for example, how well influenza vaccines protect against severe outcomes for different patient groups and how VE against influenza A(H3N2)-associated outcomes differs during seasons with and without antigenic drift. Second, all estimates are subject to residual confounding not captured by our confounders arising from potential differences in health status and risk profiles between cases and controls. For example, the patients most likely to experience severe influenza-associated outcomes (e.g., due to frailty) are also more likely to be vaccinated. Third, there may be other important outcomes not captured fully in this study, such as influenza pneumonia. Finally, while we demonstrated a clear effect on VE among patients with a high burden of comorbidities, larger and multi-season studies are necessary to determine types of comorbidities or other thresholds (e.g., ≥5 or ≥6 categories) that have the greatest impact on VE.

## Conclusions

During the 2024–2025 U.S. influenza season, characterized by sustained elevated activity, we observed significant protection from seasonal influenza vaccination against influenza-associated hospitalizations in adults overall, for both A(H3N2) and A(H1N1)pdm09 subtypes, and for all age groups, but without evidence of protection for patients with immunocompromising conditions. Substantial VE was observed against a range of severe influenza-associated outcomes, including death.

## Data Availability

All data produced in the present study are available upon reasonable request to the authors.

## Notes

^a^ See e.g., 45 C.F.R. part 46.102(l)(2), 21 C.F.R. part 56; 42 U.S.C. §241(d); 5 U.S.C. §552a; 44 U.S.C. §3501 et seq.

## Disclaimer

The findings and conclusions in this report are those of the authors and do not necessarily represent the official position of the Centers for Disease Control and Prevention (CDC).

## Statement of Potential Conflicts of Interest

I. P. reports funding from NIH and, unrelated to the current work, funding to his institution from Novartis, Regeneron, and Bluejay Diagnostics. A. L. reports prior consulting fees and research support from Roche related to baloxavir, outside of the submitted work. A. K. reports prior research funding from Dompe Pharmaceuticals, Direct Biologics and Synairgen. J. C. reports research support from Sanofi for studies influenza vaccine immunogenicity and safety, outside of the submitted work. L. B. reports receiving consulting fees from Vantive. C. G. reports consultancies for Merck and GSK and research support from NIH, CDC, AHRQ, FDA and SyneosHealth. I. V. reports funding to their institution from eMaxHealth, Evidera PPD, Vantive Health LLC, UCB Biosciences, Boehringer Ingelheim, and Eli Lilly, all outside the submitted work. M. J. reports funding from CDC for other influenza networks and studies outside of the submitted work.

## Statement of Funding

This work was supported by Centers for Disease Control and Prevention, National Center for Immunization and Respiratory Diseases (contract numbers 75D30122C12914 and 75D30122C14944 to W. H. S.).

## Notes

### Author Declarations

The Ethics Committee of U.S. Centers for Disease Control and Prevention gave ethical approval for this work. It was determined to be public health surveillance and conducted consistent with applicable federal law and CDC policy (45 CFR 46.102(l)(2)).

