## Supplemental Materials for "Influenza vaccine effectiveness against influenza A-associated hospitalization and severe in-hospital outcomes among adults in the United States, 2024–2025"

**Supplementary Materials**

Vaccine effectiveness against influenza A-associated hospitalization and severe in-hospital outcomes among adults in the United States, 2024–2025

The IVY Network

**Table of Contents**

**Appendix A**: Supplementary Methods…………………………………………………………….........1

1. Eligibility Criteria and Enrollment Practices....…….…………………………………………1
2. Severe in-hospital outcome definitions……………………………………………………..2
3. Centralized viral testing methods....…………………………………….…………………..3
4. References…………………………………………………………………………………..4

**Appendix B**: Supplementary Figures and Tables……………..……………………………………………5

**Supplementary Figure 1.** VE against influenza-associated severe in-hospital outcomes in hospitalized adults, stratified by age group; IVY Network, 2024–2025…………………………..5

**Supplementary Table 1**. Demographic and clinical characteristics of hospitalized patients vaccinated and unvaccinated against influenza, IVY Network, 2024–2025…...………………….....6

**Appendix A (Supplementary Methods):**

### 1. Eligibility Criteria and Enrollment Practices

Patients were enrolled according to the eligibility criteria listed below. Patients were enrolled as a case (influenza present), or a control (influenza absent and SARS-CoV-2 negative) based on viral testing at the time of enrollment in the hospital. Both cases and controls were hospitalized and fulfilled the same criteria for acute respiratory symptoms. Enrollment teams attempted to enroll all eligible cases and enrolled a sample of eligible controls. Controls were enrolled to maintain approximately a 1:1 case: control ratio, with controls enrolled at the same hospital as a case and within 2 weeks of case enrollment.

In addition to clinical viral testing in the local hospital, nasal swabs specimens were collected from enrolled patients and tested for influenza by RT-PCR using standardized methods at Vanderbilt University Medical Center. Patients who were originally enrolled as a control who tested positive for influenza on centralized testing were converted to cases for analysis. Thus, influenza cases in the analysis were patients hospitalized with acute respiratory symptoms who tested positive for influenza either locally in the enrolling hospital or centrally in the IVY Network central laboratory. Controls were patients hospitalized with acute respiratory symptoms who tested negative for influenza by at least one RT-PCR test, did not have any positive influenza test results during the current episode of illness, and tested negative for SARS-CoV-2.

During the period of enrollment for this influenza evaluation, cases with COVID-19 and RSV were also enrolled within the same surveillance program. The current analysis focused on influenza vaccine effectiveness was limited to influenza cases and controls. Information on the COVID-19 and RSV cases will be disseminated separately.

#### Influenza cases

<u>Inclusion Criteria for Influenza Case Enrollment:</u>

1. Age ≥18 years old.
2. Hospital admission or in an emergency department awaiting hospital admission.
3. Symptoms and/or signs compatible with an acute viral infection, including at least 1 of the following: fever; cough; shortness of breath; hypoxemia (for patients not on chronic supplemental oxygen, hypoxemia is defined as: $SpO_2$ <92% or use of supplemental oxygen to maintain $SpO_2$ ≥92%; for patients on chronic supplemental oxygen, hypoxemia is defined as $SpO_2$ below the patient's baseline $SpO_2$ or an escalation of supplemental oxygen use to maintain the baseline $SpO_2$ value); new pulmonary findings on chest imaging consistent with pneumonia.
4. Clinically obtained test that is positive for acute influenza after onset of symptoms for the current illness. The positive test may be obtained before or after hospital arrival. Examples of acute viral tests include RT-PCR tests, nucleic acid amplification tests (NAAT), and antigen tests. Serology testing may not be used for eligibility.

<u>Exclusion Criteria for Influenza Case Enrollment:</u>

1. Patient was admitted to the hospital more than 7 days ago (based on this exclusion criterion, patients must be enrolled within 7 days of hospital admission).
2. The first positive test for influenza is known to have occurred more than 10 days after onset of acute viral infection symptoms/signs listed in inclusion criterion #3. Patients with unknown onset date for COVID-19 symptoms/signs may be enrolled.
3. First positive test for acute influenza more than 3 days (72 hours) after hospital admission.
4. Any positive test for acute SARS-CoV-2 or RSV infection after symptom onset for the current illness.
5. Previously enrolled in this surveillance program within the prior 30 days.

**Controls**

Inclusion Criteria for Control Enrollment:

1. Age ≥18 years old.
2. Hospital admission or in an emergency department awaiting hospital admission.
3. Symptoms and/or signs that overlap with an acute viral respiratory infection, including at least 1 of the following: fever; cough; shortness of breath; hypoxemia (for patients not on chronic supplemental oxygen, hypoxemia is defined as: $SpO_2$ <92% or use of supplemental oxygen to maintain $SpO_2$ ≥92%; for patients on chronic supplemental oxygen, hypoxemia is defined as $SpO_2$ below the patient's baseline $SpO_2$ or an escalation of supplemental oxygen use to maintain the baseline $SpO_2$ value); new pulmonary findings on chest imaging consistent with pneumonia.
4. Clinically obtained test that is negative for acute SARS-CoV-2, influenza, or RSV after onset of symptoms for the current illness. The negative test may be obtained before or after hospital arrival. Examples of acute SARS-CoV-2 tests include RT-PCR tests, NAAT, and antigen tests. Serology testing may not be used for eligibility.

Exclusion Criteria for Control Enrollment:

1. Patient was admitted to the hospital more than 7 days ago (based on this exclusion criterion, patients must be enrolled within 7 days of hospital admission).
2. The first negative test for acute SARS-CoV-2, influenza or RSV infection is known to have occurred more than 10 days after onset of symptoms/signs listed in inclusion criterion #3. Patients with unknown onset date for symptoms/signs may be enrolled.
3. First negative test for acute viral infection (SARS-CoV-2, influenza, or RSV) more than 3 days (72 hours) after hospital admission.
4. Any positive test for acute SARS-CoV-2 (≥18 years), RSV (≥60 years), or influenza infection after symptom onset for the current illness.
5. Previously enrolled in this surveillance program within the prior 30 days.
6. Inability to obtain an upper respiratory sample for central laboratory testing within 10 days of symptom onset for the current illness.

**2. Severe in-hospital outcome definitions**

The influenza vaccine effectiveness analysis included the following five influenza-associated severe in-hospital outcomes:

a) Standard oxygen therapy
b) Non-invasive advanced respiratory support
c) Invasive organ support
d) ICU admission
e) Death

Patients who were receiving chronic organ support prior to influenza infection were not eligible for the invasive organ support outcome for that organ system. For example, patients who were receiving chronic renal replacement therapy prior to influenza infection were not eligible for the renal replacement therapy outcome. Additionally, patients were only included in a respiratory outcome if the level of support received in the hospital exceeded the baseline level of oxygen support. A patient receiving non-invasive ventilation at home prior to influenza infection, for example, would not qualify for the standard oxygen therapy or non-invasive advanced respiratory support outcomes but would qualify for the invasive organ support outcome if they received invasive mechanical ventilation in-hospital.

**a) Standard oxygen therapy:**

Influenza-associated hospitalization plus new receipt of standard flow supplemental oxygen (<30 liters/min) within 28 days of hospitalization.

**b) Non-invasive advanced respiratory support:**

Influenza-associated hospitalization plus new receipt of high-flow nasal cannula (≥30 liters/min) or non-invasive ventilation (continuous positive airway pressure [CPAP] and bilevel positive airway pressure [BiPAP] delivered through mask as therapy for acute illness) within 28 days of hospitalization.

**c) Invasive organ support:**

Influenza-associated hospitalization plus new invasive mechanical ventilation (positive pressure administered through an endotracheal tube or tracheostomy tube), renal replacement therapy, or intravenous vasopressor medication use (norepinephrine, epinephrine, dopamine, phenylephrine, and vasopressin) within 28 days of hospitalization.

**d) Intensive care unit (ICU) admission:**

Influenza-associated hospitalization plus ICU admission within 28 days of hospitalization.

**e) Death:**

Influenza-associated hospitalized plus death within 28 days of hospitalization.

### 3. Centralized viral testing methods

Personnel at each site collected a nasal swab specimen by either a fresh swabbing procedure or from a residual aliquot in the clinical laboratory at enrollment and sent them to Vanderbilt University Medical Center (VUMC; Nashville, TN). Overnight shipments of specimens were received on dry ice and maintained at −80°C for ≤14 days, thawed, and aliquoted into 2-mL cryovials. Aliquoted material was immediately returned to −80°C for long-term storage. At the time of thawing, a volume of specimen sufficient for pathogen testing was combined with lysis buffer and held at 4°C for <1 hour, followed by automated nucleic acid extraction. Specimens were received at the central laboratory in original containment. Research specimens were collected in 3 mL viral transport medium and processed at VUMC. Salvaged clinical residuals were received in containers used by the local site diagnostic laboratory and further processed (i.e., aliquoted) at VUMC, where they underwent testing for influenza virus, SARS-CoV-2, and RSV by RT-PCR. For specimens that tested positive for influenza virus by RT-PCR, an aliquot of primary specimen stored at −80°C was shipped overnight on dry ice to University of Michigan (Ann Arbor, MI) for viral whole-genome sequencing (WGS). More detailed methods used by IVY can be found previously published from the 2021–2022 and 2022–2023 seasons.[1,2]

### Appendix B (Supplementary Figures and Tables):

| Group | Vaccinated / Total (%) | | VE % (95% CI) |
|---|---|---|---|
| | Cases | Controls | |
| **Standard oxygen therapy** | | | |
| 18–64 years | 96 / 368 (26) | 726 / 2128 (34) | 39 (21–53) |
| ≥65 years | 171 / 406 (42) | 1203 / 2246 (54) | 43 (28–54) |
| **Non-invasive advanced respiratory support** | | | |
| 18–64 years | 28 / 104 (27) | 726 / 2128 (34) | 32 (-7–57) |
| ≥65 years | 46 / 126 (37) | 1203 / 2246 (54) | 53 (32–67)[a] |
| **Invasive organ support** | | | |
| 18–64 years | 26 / 159 (16) | 726 / 2128 (34) | 65 (45–77) |
| ≥65 years | 44 / 118 (37) | 1203 / 2246 (54) | 53 (31–68) |
| **ICU admission** | | | |
| 18–64 years | 32 / 220 (15) | 726 / 2128 (34) | 69 (54–79) |
| ≥65 years | 75 / 194 (39) | 1203 / 2246 (54) | 50 (32–63) |
| **Death** | | | |
| 18–64 years | 4 / 24 (17) | 726 / 2128 (34) | 64 (9–86)[a] |
| ≥65 years | 15 / 36 (42) | 1203 / 2246 (54) | 43 (-7–69)[a] |

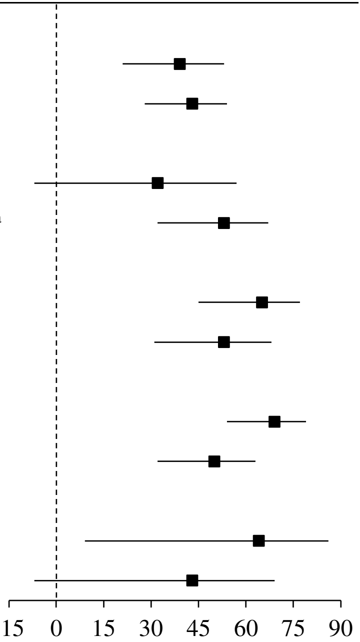


Abbreviations: IVY = Investigating Respiratory Viruses in the Acutely Ill; VE = vaccine effectiveness; CI = confidence interval.

[a]Firth regression used due to separation.

**Supplementary Figure 1.** VE against influenza-associated severe in-hospital outcomes in hospitalized adults, stratified by age group; IVY Network, 2024–2025.

**Supplementary Table 1.** Demographic and clinical characteristics of hospitalized patients vaccinated and unvaccinated against influenza, IVY Network, 2024–2025.

| | **Overall (n = 6109)** | **Vaccinated (n = 2511)** | **Unvaccinated (n = 3598)** | ***p*-value** |
|---|---|---|---|---|
| **Demographic characteristics** | | | | |
| Census region | | | | 0.15 |
| Northeast | 1565 (26) | 621 (25) | 944 (26) | |
| Midwest | 1354 (22) | 539 (21) | 815 (23) | |
| South | 1381 (23) | 571 (23) | 810 (23) | |
| West | 1809 (30) | 780 (31) | 1029 (29) | |
| Age, years (median [IQR]) | 65 [53–75] | 69 [59–78] | 62 [49–72] | **<0.01** |
| Age group | | | | **<0.01** |
| 18–49 years | 1242 (20) | 319 (13) | 923 (26) | |
| 50–64 years | 1749 (29) | 612 (24) | 1137 (32) | |
| ≥65 years | 3118 (51) | 1580 (63) | 1538 (43) | |
| Female | 3146 (51) | 1310 (52) | 1836 (51) | 0.39 |
| Race/ethnicity | | | | **<0.01** |
| White, non-Hispanic | 3374 (55) | 1538 (61) | 1836 (51) | |
| Black, non-Hispanic | 1425 (23) | 496 (20) | 929 (26) | |
| Hispanic, any race | 823 (13) | 286 (11) | 537 (15) | |
| Other race, non-Hispanic[a] | 330 (5) | 147 (6) | 183 (5) | |
| Other[b] | 157 (3) | 44 (2) | 113 (3) | |
| **Patient characteristics** | | | | |
| Influenza case | 1735 (28) | 582 (23) | 1153 (32) | **<0.01** |
| Health status indicators | | | | |
| Insured | 5917 (97) | 2485 (99) | 3432 (95) | **<0.01** |
| Current tobacco smoking | 1297/5905 (22) | 357/2460 (15) | 940/3445 (27) | **<0.01** |
| ≥1 hospitalization in prior year | 3744 (61) | 1652 (66) | 2092 (58) | **<0.01** |
| Chronic respiratory condition | 2642 (43) | 1190 (47) | 1452 (40) | **<0.01** |
| Immunocompromised[c] | 1621 (27) | 752 (30) | 869 (24) | **<0.01** |
| Number of chronic underlying medical conditions[d] | 2 [1–3] | 3 [2–3] | 2 [1–3] | **<0.01** |
| **Admission characteristics** | | | | |
| Illness onset to influenza testing, days (median [IQR]) | 2 [0–4] | 2 [0–3] | 2 [0–4] | 0.25 |
| Illness onset to hospitalization, days (median [IQR]) | 2 [0–4] | 2 [0–3] | 2 [0–4] | 0.62 |
| **Severe in-hospital outcomes[e]** | | | | |
| Standard oxygen therapy | 774/1692 (46) | 267/562 (48) | 507/1130 (45) | 0.33 |
| Non-invasive advanced respiratory support | 284/1692 (17) | 102/562 (18) | 182/1130 (16) | 0.32 |
| Invasive organ support | 277/1692 (16) | 70/562 (12) | 207/1130 (18) | **<0.01** |
| ICU admission | 414/1692 (24) | 107/562 (19) | 307/1130 (27) | **<0.01** |
| Death | 60/1735 (3) | 19/582 (3) | 41/1153 (4) | 0.86 |

Abbreviations: IVY = Investigating Respiratory Viruses in the Acutely Ill; IQR = interquartile range.

[a]"Other race, non-Hispanic" includes non-Hispanic Asian, Native American or Alaska Native, and Native Hawaiian or other Pacific Islander; these groups were combined due to small counts.
[b]"Other" includes patients who self-reported their race and ethnicity as "Other" and those for whom race and ethnicity were unknown.

[c]Immunocompromising conditions included active solid tumor or hematologic cancer, solid organ transplant, bone marrow/stem cell transplant, HIV infection, congenital immunodeficiency syndrome, use of an immunosuppressive medication in the past 30 days, splenectomy, or another condition that causes moderate or severe immunosuppression.
[d]Chronic underlying medical conditions included cardiovascular disease, neurologic disease, pulmonary disease, gastrointestinal disease, endocrine disease, renal disease, hematologic disease, and autoimmune/inflammatory disease.
[e]Data on severe in-hospital outcomes was not collected for control patients. Patients are only included in the outcome if the outcome is an elevation in severity from their baseline.
